# Predominance of Den 2 and Den 3 serotypes during the 2025 dengue outbreak in Chattogram, Bangladesh: Implications for Public Health Preparedness

**DOI:** 10.64898/2026.02.21.26346763

**Authors:** Rajat Sanker Roy Biswas, Tamal Moharar, Mohammad Rezaul Karim, Md. Mahbub Hasan, Sanjoy Kanti Biswas

**Affiliations:** Department of Medicine, Chattogram Maa O Shishu Hospital Medical College, Agrabad, Chattogram, Bangladesh; Department of Microbiology and Molecular Biology, Chattogram Maa O Shishu Hospital Medical College; Department of Medicine, Chattogram Medical College; Department of Genetic Engineering and Biotechnology, Faculty of Biological Sciences,University of Chittagong, Chattogram 4331, Bangladesh; Department of Microbiology and Molecular Biology, Chattogram Maa O Shishu Hospital Medical College, Chattogram, Bangladesh

**Keywords:** Dengue, Serotype, RT-PCR, Outbreak, Bangladesh

## Abstract

**Introduction:** Dengue has been prevalent in a regular fashion in Bangladesh and Chattogram for the last 6-7 years and is showing some serotype twisting. So, the objectives of the present study were to explore the burden of dengue serotypes in Chattogram.

**Methods:** In this study, 223 Dengue RT-PCR positive patients were evaluated for serotyping. Gender and age group, along with cycle threshold (CT) values, were also collected. Data after collection were compiled, analyzed, and plotted in Microsoft Excel and GraphPad Prism 10.4. Ethical clearance was taken to conduct the study.

**Results:** Among 223 patients analyzed, males and females were found near equal (113 and 110). Middle-aged patients were more than the extremes of age. The mean ± SD of age was 33.55 ± 13.67 years. Regarding serotype distributions, isolated Den 1, Den 2 and Den 3 were found 1.3%, 73.1% and 6.7%, respectively. Concurrent infections with multiple serotypes were observed in several patients, most notably the Den 2 and Den 3 combination, which accounted for 14.3% (n=32) of the cases. Other co-infections were less frequent: the Den 1 and Den 2 pairing appeared in 3.6% (n=8) of the cohort, while triple-serotype infections (Den 1, 2, and 3) and Den 3/Den 4 pairings were rare, each occurring in only 0.4% of patients. Statistical analysis of CT values revealed no significant sex-based differences for Den 2 and Den 3. However, significant variations in CT values were observed when comparing Den 1 against both Den 2 and Den 3 (p < 0.05). In contrast, the difference between Den 2 and Den 3 Ct values remained statistically insignificant.

**Conclusion:** In the year 2025, Dengue serotypes 2 and 3 were found to be the most prevalent, both in isolated or in combinations and Den 1 and Den 4 were found minimum. Exposure to multiple serotypes and twisting from one serotype to another might influence the dengue outcome in future, which needs further exploration.

## Introduction

Dengue is a viral disease which is transmitted by mosquitoes of the Aedes genus, mainly by *Aedes aegypti* and sometimes by *Aedes albopictus* and by some other rare species. Dengue is considered a major global public health issue as it is associated with mortality and morbidity worldwide. In recent years, the incidence of dengue increased globally, though the majority of cases are asymptomatic or present with mild symptoms. However, a good number of patients progress to severe forms, and those patients needed hospitalization and can result in death, mainly those who are vulnerable group like patients of extremes of ages, pregnant women and patients with multiple comorbidities.[1]

Bangladesh is currently experiencing a countrywide dengue outbreak, marked by an expansion of the disease into a year-round threat that now saturates all 64 districts. While the country faced its deadliest outbreak in 2023 with 1,705 deaths, recent data from early 2025 suggests continued transmission even during traditionally low-risk months.[2]

In the year 2025, roughly 105,562 cases and 412 deaths were recorded, indicating a decrease in mortality compared to previous years, but a persistent high volume of infections. There were 101,214 cases and 575 deaths recorded throughout 2024[2]

Dengue virus, which is an Arbobovirus under Flavivirus genus and it has four serotypes like Den −1 to Den −4. There are different genotypes of all four serotypes, also. Dengue clinical features and presentations, along with its severity, are related to the type and number of serotype involve. Multiple serotype infection at the same time in the same patient causes a worse outcome[3]

Infection with one of the four serotypes induces lifelong specific immunity against that serotype, but does not protect against the other serotypes. This can favor the occurrence of severe dengue in a second infection due to immunopathological phenomena such as antibody-dependent enhancement.[4] The co-circulation of several serotypes in the same region has been associated with an increase in the severity of cases and large-scale outbreaks [5,6].

In Bangladesh, serotype Den 2 has been prevailing for the last two to three years, but shifting of serotypes is predicted, though study is rare in the context of Chittagong, Bangladesh. The objectives of the present study were to document the types of serotypes prevailing in the year 2025 in Chattogram, Bangladesh.

## Methods

The present study was conducted at the Department of Microbiology and Medicine of Chattogram Maa O Shishu Hospital Medical College (CMOSHMC), Chattogram, Bangladesh, during a six-month study period from 01/07/2025 to 31/31/2025 223. The study strictly followed to protect the privacy, anonymity, and rights of every participant. The study team fulfilled ethical approval requirements from the institutional review board of the research administration of CMOSHMC for the serotype analysis of every sample.

Samples from the patients, who were aged between 5 and 65 years with clinical symptoms of dengue (e.g., onset of fever between 2 and 5 days and/or other symptoms, i.e., rash, myalgia, bone pain, headache, nausea, vomiting, and diarrhea, etc.), were initially evaluated in this study from attending inpatients and outpatient department of the hospital. Those who were severely ill or who had any acute condition that suggested that invasive sample collection should be avoided were excluded from this study.

With all aseptic and universal precautions, 3mL of venous blood was collected in a serum-separating tube (SST) from each febrile patient included. Serum samples were separated by centrifugation from SSTs and subsequently aliquoted into 1.5-mL Eppendorf tubes. Every sample had been subjected to primary screening by a dengue NS1 rapid diagnostic test (RDT) using the Bioline dengue NS1 antigen kit (Abbott, Ingbert, Germany).

Among the NS1 positive cases, RNA was extracted from 150mL of serum by use of CE IVDR Marked GeneProof RNA Extraction Kit; Czeck Republic according to the manufacturer’s instructions. Each RNA sample was cryopreserved at –80_C until batch wise real-time reverse transcription polymerase chain reaction (RT-PCR) for serotype detection was carried out. Dengue virus RNA was detected using the CDC-recommended in-vitro qualitative RT-PCR assay of Den 1 to Den 4 by RUO Marked NeoDx Dengue Genotyping Real-Time PCR Kit; India. An amplification curve with a cycle threshold (Ct) of <38 was evaluated as positive, and Ct values of >38 were evaluated as negative.[6]

A total of 223 patients were evaluated serologically, and those were finally included in the study. Gender and age group were collected, and CT values for different dengue serotypes were also collected. Data after collection was compiled and analyzed and plotted by Microsoft Excel and GraphPad Prism 10.4. CT values for Den 2 and Den 3 in relation to gender differences were evaluated by a violin plot. Also, CT values of Den 1 and Den 2, Den 1 and Den 3, and Den 2 and Den 3 were compared. The Mann-Whitney U test was employed to compare the median CT values between male and female cohorts, as the data did not follow a normal distribution. Ethical clearance was taken to conduct the study from the ethical review board of the medical college (CMOSHMC/IRB/2025/05).

## Results

Among 223 patients analyzed for serotyping, males and females were found to be nearly equally distributed (113 and 110) [Figure 1]. Age group distribution revealed, <12 years patients were 3(1.3%), 13-20 years were 37(16.6%), 21-30 years were 70(31.4%), 31 – 40 years were 55(24.7%), 41-50 years 29(13.0%), 51-60 years were 19(8.5%) and > 61 years were 10(4.5%). Mean± SD of age was found to be 33.55 ± 13.67 years [Figure 2]. Regarding serotype distributions of study patients, isolated Den 1, Den 2 and Den 3 were found 1.3%, 73.1% and 6.7%, respectively. Combined Den 1 and Den 2 were 8(3.6%), Den 2 and Den 3 were 32(14.3%), Den 1, Den 2 and Den 3 were 1(0.4%) and Den 3 and Den 4 were 1(0.4%) [Table 1]. CT values of Den 1, Den 2, Den 3 and Den 4 were calculated and presented in a Violin plot [Figure 3]. CT values of male and female patients positive for Den 2 and Den 3 were found not to be significant [Figure 4].

**Table 1:**
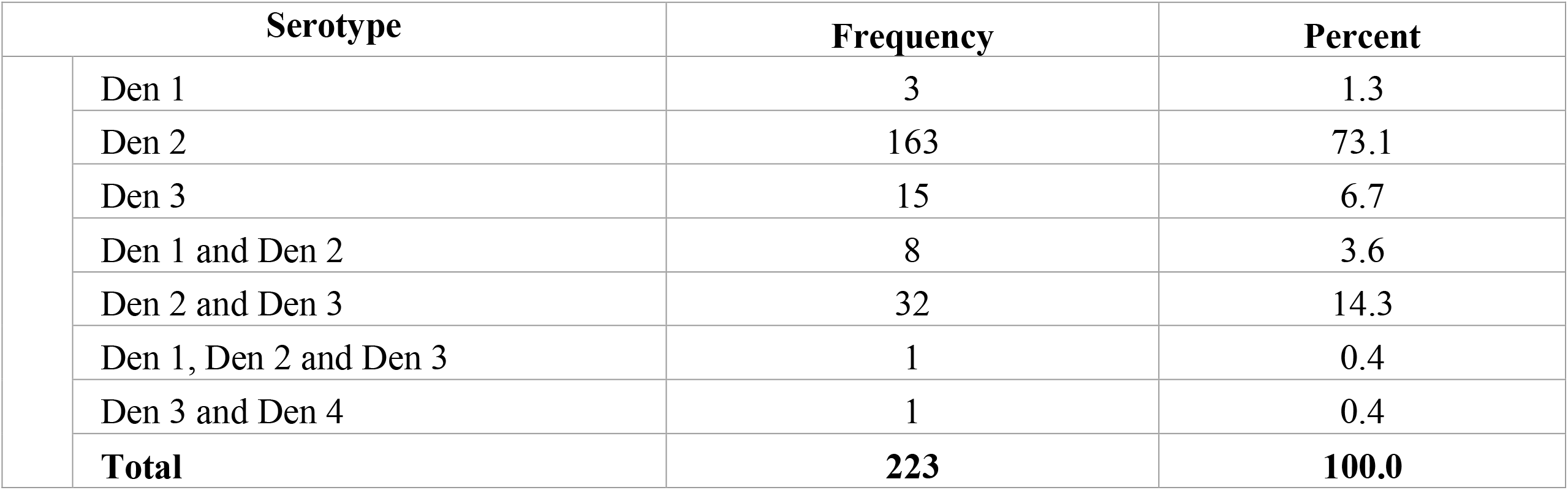
Distribution of Dengue Virus (Den) serotypes and co-infections among the study population (N=223).

**Figure 1.**
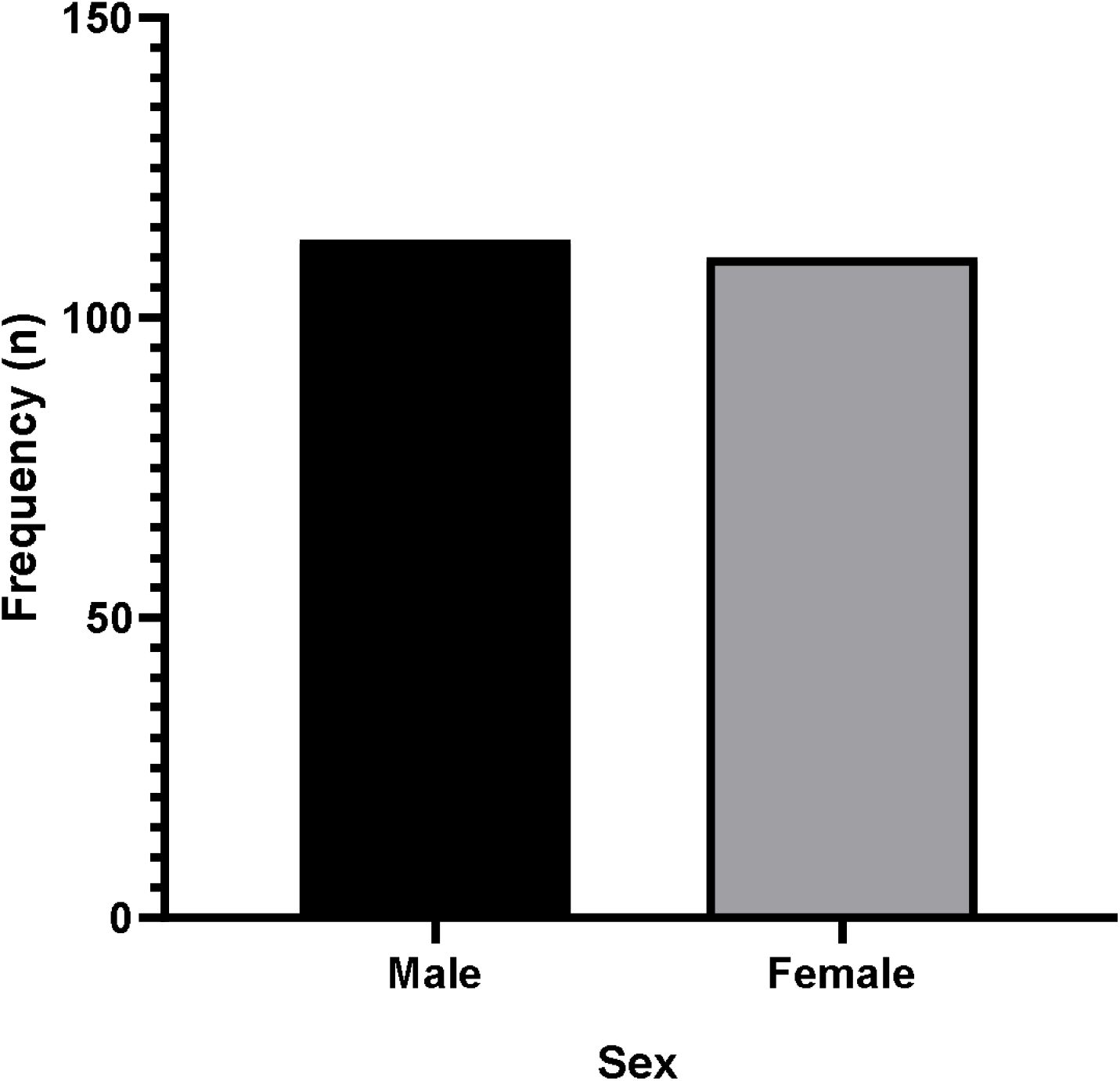
Distributions of the study participants by sex (N=223).

**Figure 2.**
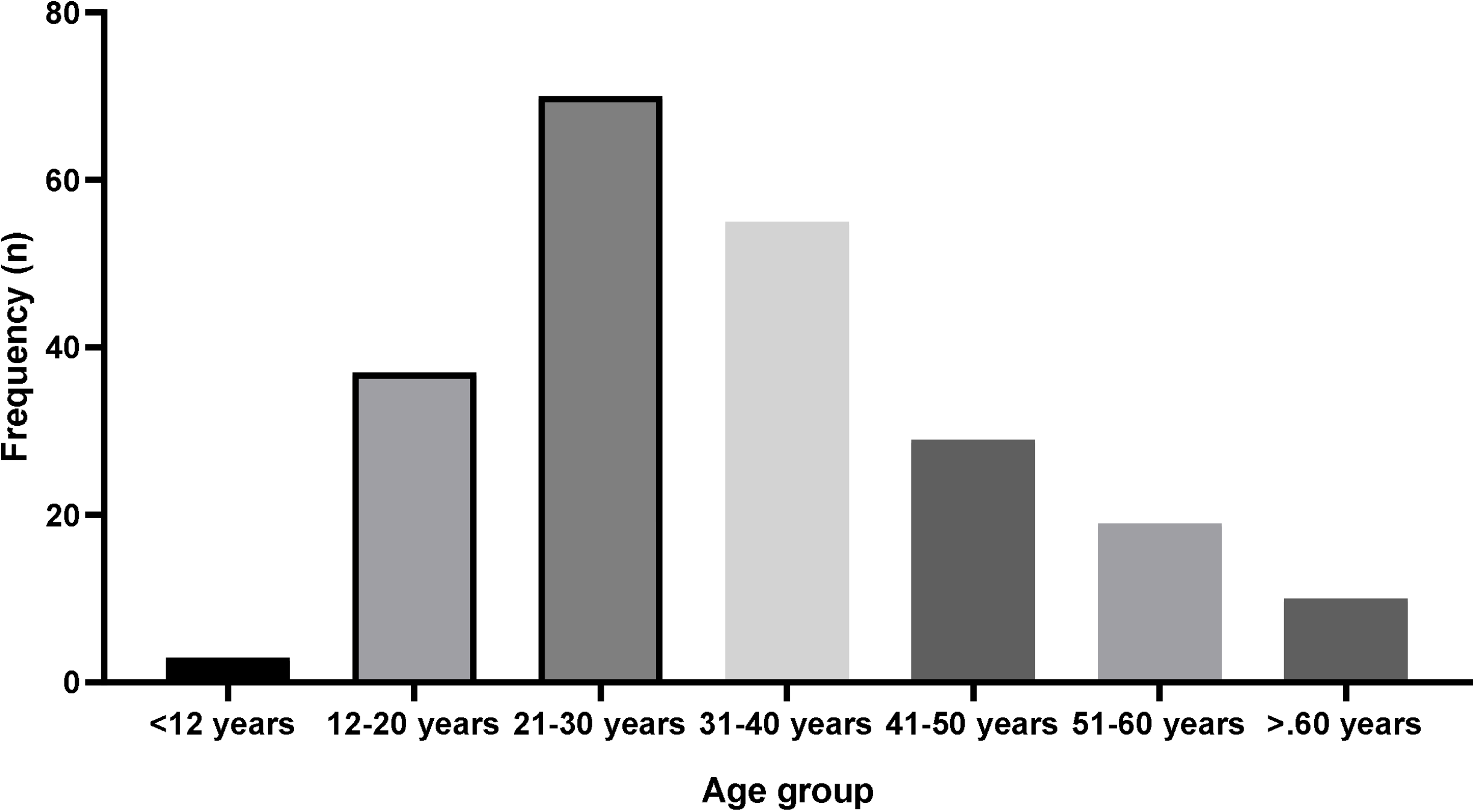
Distributions of dengue cases across different age groups (N=223).

**Figure 3.**
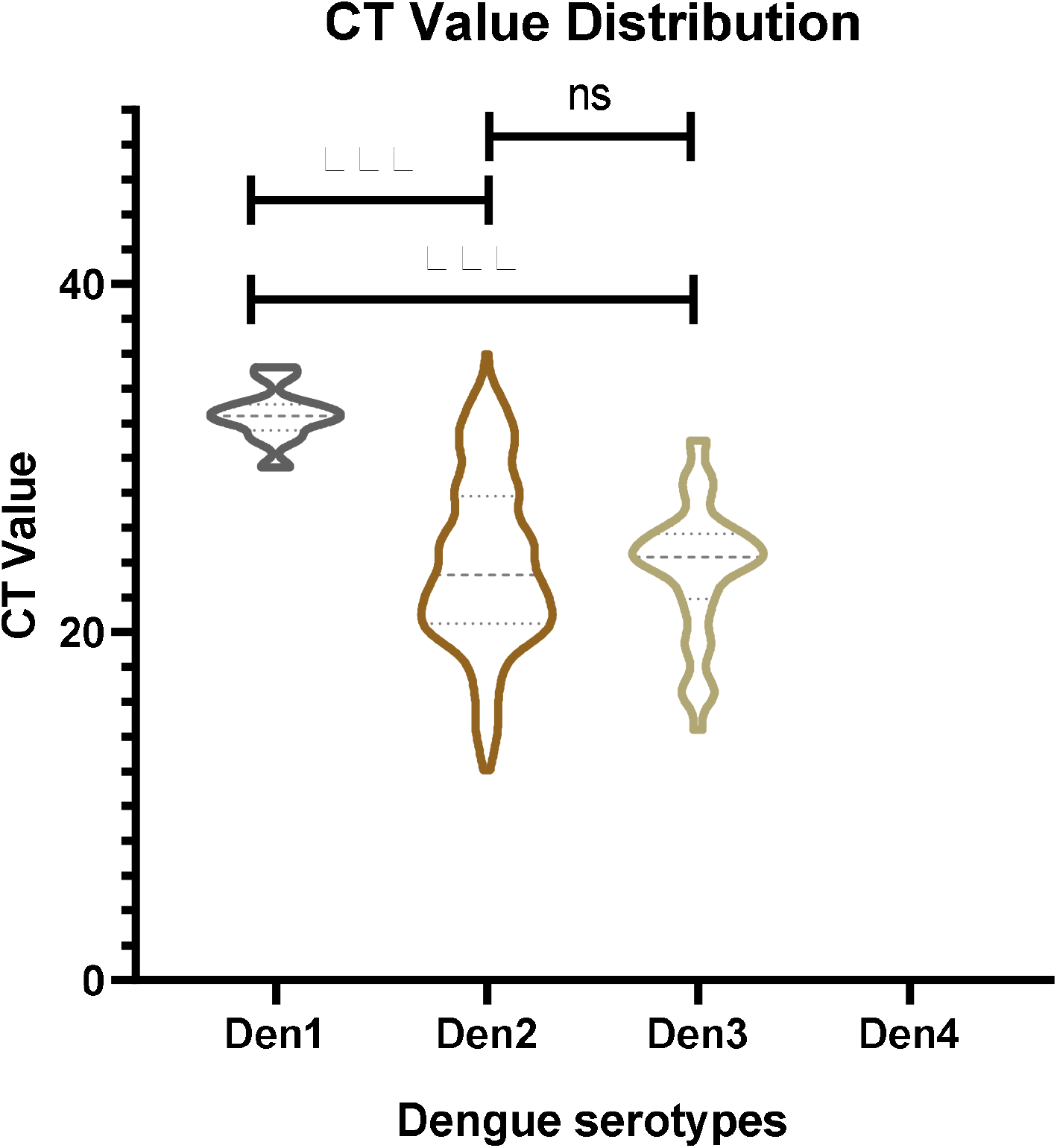
Cycle threshold values (CT) across different Den serotypes. The violin plot illustrates the distribution and density of CT values for each serotype. Statistical significance was determined via the Mann-Whitney U test (*** p < 0.001; ns: not significant).

**Figure 4.**
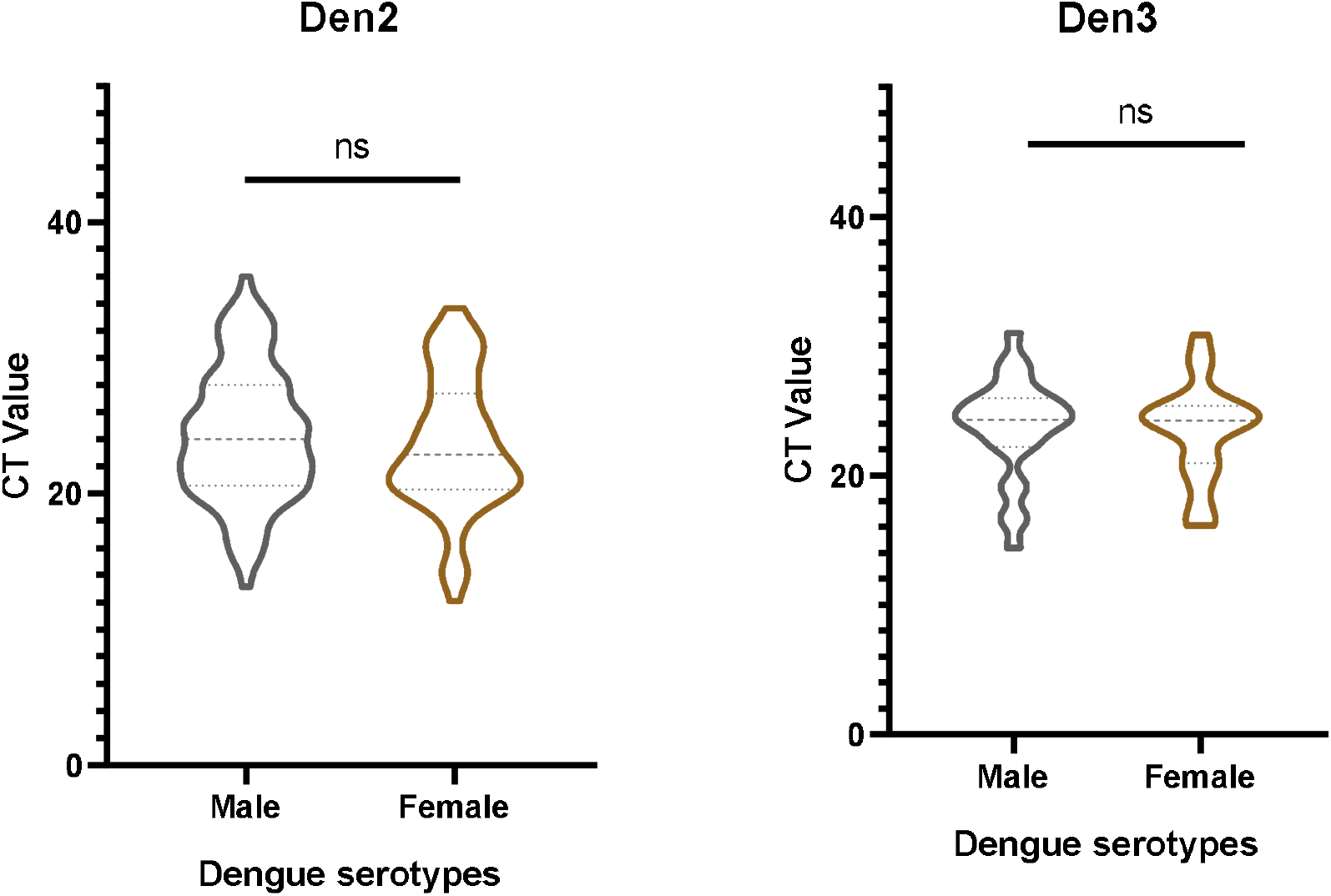
Comparison of Cycle threshold (CT) values by sex for Den 2 and Den 3. Violin plots represent the distribution of viral loads between male and female patients for the two most prevalent serotypes. No statistically significant difference in CT values was observed between sexes for either serotype (Mann-Whitney U test; ns = not significant).

## Discussion

The present study was conducted among 223 RT PCR positive dengue patients, where the main serotypes prevailing in Chattogram were Den 2(73.1%) and Den 3(6.7%). Again, a significant number of patients were infected with combined Den 1 and Den 2-8(3.6%), Den 2 and Den 3 −32(14.3%). Co-infections were also found, though they were very low in number. In the years 2023 and 2024, Den 2 prevailed the maximum, but in the year 2025, there are shifting of Den 3 along with Den 2 and a good number of patients were found infected with Den 3 along with Den 2 combined. However, until 2012, there were few data available regarding circulating serotypes of Den. Between 2013 and 2018, Den-2 was detected as the dominant serotype, followed by a moderate number of Den1 cases and a few cases of Den-3.[8] That pattern was changed by the reemergence of Den-3 as the predominant serotype in Dhaka City in subsequent years (2019–2022)[9,10]

Shifting and twisting of serotypes may be related to mortality and morbidity. These heterogenicity of serotypes, if prevailing in the coming years, might cause a greater number of antigenic reactions and might increase the morbidity and mortality.[7] It was established that prior antibodies produced by one serotype exposure are unable to confer protection against another serotype. Again secondary infection with heterotypic serotype is frequently associated with severe abd fatal clinical menifestations which is likely related to antibody-dependent enhancement.[11] So we have to prepare for facing such conditions, and the authority should take action regarding mosquito control and destroying the breeding places.

In the present study, male and female patients were affected in a near same number, and the affected age groups were mainly younger. A study done previously [10] found that males were more than females (77.03% vs 22.97%), and younger age groups were more vulnerable, which was from 21 years to 40 years.

Another study done earlier found the majority of the patients belonged to the age group (26-40 years), and the mean (36.44 years) aligns with other relevant studies done in the sub-continent region reporting a predominance of young to middle-aged adults with dengue infection.[12,13] This could be partially explained by behavioural differences, such as males spending more time outdoors for work or leisure activities, potentially increasing their exposure to mosquito bites. However, further investigation is necessary to understand the interplay of social norms, occupational risks, and biological susceptibility in this context.

An efficient and comprehensive vector control and community education strategy can help to limit the mosquito flow from the country. Upgrading the knowledge of health care providers across the country should be implemented by the leading authority of the country. Development of a tetravalent vaccine is a dire need to control the immune-enhanced severity. For that local and global collaboration is urgently needed.

In conclusion, we can say that dengue may prevail for a long time in Bangladesh, as we are still not ready to control the vector. Serotype twisting might influence its severity and outcome, though the present study did not analyze the outcome of the included patients, which is a limitation of the study. Social leaders and health care professionals should make themselves ready to combat these diseases both in the community setting and at the health care delivery level. Large scale countrywide analysis is needed to get the country’s scenario.

## Data Availability

Data are available after valid request

## Author contribution

RSRB-Protocol design, fund collection, methodology, data collection, report writing and final report, TM-Data collection and laboratory works, MRK-Protocol preparation and report writing, MMH-Data analysis, report writing and final report editing, SKB-Fund collection and final report editing

